# Reduced dietary protein intake does not alter autophagy in human blood: a randomized crossover study in healthy adults

**DOI:** 10.64898/2026.07.07.26357503

**Authors:** Sanjna Singh, Célia Fourrier, Leanne K. Hein, Julien Bensalem, Alexis Martin, Kathryn J. Hattersley, Barbara King, Xiao Tong Teong, Kathy Baker, Kylie Lange, Gemma Barker, Jemima R. Gore, Leonie K. Heilbronn, Timothy J. Sargeant

## Abstract

**Background & aims:** Autophagy activation is a promising strategy to counteract age-related cellular dysfunction. While preclinical studies suggest dietary protein restriction can induce autophagy via mTORC1 inhibition, direct human evidence using dynamic, flux-based measurements remains limited. The aim of this study was to determine whether a low protein diet could modulate autophagic flux in humans.

**Methods:** We conducted a randomized crossover trial in which 74 healthy adults were randomized to receive two 4-week interventions of either average-protein (20% energy) or reduced-protein (10% energy) diets prescribed to maintain calculated energy balance, separated by a 4-week washout period. The primary outcome was autophagic flux measured in whole blood using a validated assay that preserves PBMCs in their physiological environment during lysosomal inhibition. Secondary outcomes included metabolic markers, body composition, and self-reported health metrics.

**Results:** Sixty-three participants completed both interventions (mean ± SD age 29.5 ± 7.2 yrs; BMI 24.0 ± 3.2 kg/m^2^). Reducing protein intake did not alter autophagic flux (adjusted mean difference: −8.46 ng LC3B-II/mg protein/h; 95% CI: −24.06 to 7.14; p = 0.28). Metabolomic profiling confirmed effective dietary separation, with lower circulating urea following reduced protein intake. Small differences in body weight and muscle mass were observed, while fat mass was unaffected. Fasting glucose, insulin, lipids, blood pressure, and quality of life did not differ between the two diets.

**Conclusions:** Moderate protein restriction does not increase basal autophagy in circulating immune cells of healthy adults, suggesting protein reduction alone, without caloric deficit, may be insufficient to activate autophagy in human blood.

**Clinical trial registry number:** Australian New Zealand Clinical Trials Registry Identifier ACTRN12623000260628 https://anzctr.org.au/Trial/Registration/TrialReview.aspx?id=382790

## Introduction

Autophagy is a repair and recycling pathway that is activated in response to cellular stress – such as during nutrient scarcity, or in the presence of damaged organelles or misfolded proteins – to sequester material and subsequently deliver it to the lysosome for breakdown. Defects in this process have been extensively linked to age-related pathologies such as neurodegeneration (1) and cardiometabolic disease (2). In fact, dysfunctional autophagy itself is a hallmark of aging (3), although there is emerging evidence to suggest that it does not decline uniformly in all contexts or across all tissues (4–8). Further, several of the health and longevity benefits of calorie restriction have been attributed, at least partly, to autophagy (9). Consequently, activation of autophagy is of immense interest to the aging field, as it has the potential to ameliorate age-related damage to improve life- and health-span.

Limiting nutrient intake, for instance through calorie restriction, is a well-established inducer of autophagy, and acts via inhibition of mechanistic target of rapamycin complex 1 (mTORC1), a key autophagy repressor. However, energy restriction is not always suitable as adherence over long time periods can be difficult and may carry potential risks. Reducing protein intake may provide an alternate nutritional strategy, as it has also been shown to reduce mTORC1 activity in the liver (10) and brain (11) and increase autophagic markers across various tissues in rodents (12, 13). Further, increased carbohydrate: protein ratios are associated with increased lifespan in animals (10) and are correlated with reduced risk of death in people aged 50-65 and reduced risk of cancer and diabetes mortality (14). Complementing these findings, high-protein diets conversely activate macrophage mTORC1 signalling and impair lysosome and autophagy-related processes in ways that exacerbate cardiovascular disease phenotypes in preclinical models (15, 16). Together, these data suggest that dietary protein manipulation may represent a feasible strategy to modulate autophagy in humans, but direct clinical evidence, particularly using measures of autophagic ‘flux’ – which reflects the actual turnover of the pathway rather than just a steady-state snapshot measurement – is limited (17, 18).

In humans, evidence linking dietary protein intake to autophagy is sparse and is derived predominantly from short-term dietary challenges. Although a single high protein meal activates mTORC1 signalling in blood circulating immune cells (peripheral blood mononuclear cells, PBMCs), its effects on autophagy are unclear: LC3-positive puncta decrease in monocytes (15) yet autophagic flux, which reflects the actual turnover of this pathway, in the total PBMC population appears unchanged (19). Critically, whether sustained reductions in dietary protein intake can modulate autophagic flux in humans has not been directly tested. Circulating immune cells play central roles in age-related inflammation and cardiometabolic disease (20, 21), making blood-based measures of autophagy particularly relevant to human aging. In this randomized crossover trial, we sought to determine whether a 4-week-long reduction of dietary protein alters autophagic flux in comparison to standard protein intake. To do this, we employed an assay which measures autophagic flux via turnover of the autophagic marker LC3B in whole blood (22), thus preserving PBMCs in their physiological environment. Secondary outcomes included assessment of metabolic and physiological adaptations to the dietary intervention.

## Materials & Methods

### Study design and participants

This single-site, single-blind, randomized crossover trial was approved by the University of Adelaide Human Research Ethics Committee (H-2021-154) and prospectively registered with the Australian New Zealand Clinical Trials Registry ACTRN12623000260628 (23). Healthy adults aged between 20 and 50 years of age, with a body mass index (BMI) between 18.5 and 29.9 kg/m^2^ were recruited from the Adelaide metropolitan area between March 2023 and June 2024. Key exclusion criteria were co-morbidities likely to affect lysosomal system activity (e.g. cancer, cardiovascular disease, diabetes); use of medications known to impact autophagy, appetite, body composition or metabolism (e.g. anti-inflammatory or blood glucose lowering medications); self-reported alcohol or substance abuse; smoking; allergies or dietary restrictions incompatible with the study intervention; recent weight fluctuations (>5% weight gain or loss in the last 3 months); high consumption of protein or supplements; dietary practices that may, in the opinion of the principal investigator, affect autophagy (e.g. calorie restriction, intermittent fasting). The criteria also excluded women who were pregnant, breastfeeding, planning a pregnancy, peri- or post-menopausal.

Participants were randomized 1:1 to one of two dietary sequences (average-protein followed by reduced-protein, or vice versa) using a computer-generated randomization schedule using randomly permuted blocks of size 4 and stratified by sex. Each dietary period lasted 4 weeks and was separated by a 4-week washout period. The washout was considered sufficient to prevent carry-over of dietary effects from the first to the second period.

Given the nature of the intervention, the investigator responsible for participant interviews and diet provision was not blinded to allocation; however, all laboratory analyses, data management, and statistical analyses of primary outcomes were conducted by investigators blinded to group assignment. Participants were also blinded to dietary condition, with study aims described in general terms and diets matched as closely as possible in appearance and composition to minimise expectancy bias.

### Diet interventions

Study diets were designed by a research dietitian using FoodWorks Professional (version 10, Xyris Software Australia). Participants received individualised 7-day rotating menus tailored to personal preferences (e.g. cultural preferences, food intolerances, vegetarian), with most foods supplied by the study. To facilitate masking and maintain dietary similarity, macronutrient targets were achieved using commercially available high- or low-protein supplemental powders, repackaged into identical unlabelled sachets. As supplements were dairy-based, lactose intolerance was an exclusion criterion. Participants were asked to complete daily food checklists and document any deviations from the prescribed intake.

#### Average Protein Diet (Control)

The control diet was delivered at calculated energy balance based on previously validated equations (24). Its macronutrient profile comprised 20% protein, 35% fat (<10% saturated), and 45% carbohydrate, with >30 g of fibre daily.

#### Reduced Protein Diet (Intervention)

The intervention diet closely matched the control, differing only in macronutrient distribution: 10% protein, 35% fat (<10% saturated), and 55% carbohydrate, with >30 g of fibre daily.

### Outcome measures

This study aimed to determine whether reducing dietary protein intake in healthy adults could increase autophagy. The primary outcome was change in autophagic flux between diets. Secondary outcomes included changes in blood metabolic markers (fasting plasma glucose, lipids and insulin), body mass and composition, muscular strength, and self-reported sleep quality, physical activity, eating behaviours, diet satisfaction and health-related quality of life.

#### Blood collection

Participants attended four study visits: at the beginning and end of each 4-week dietary intervention period. Visits were conducted in the morning following an overnight fast (≥12 h), with abstinence from alcohol, caffeine, and strenuous exercise. Blood samples were collected by venipuncture and processed within 30 minutes.

#### Anthropometry, blood pressure and grip strength

During each study visit, body weight, height, waist and hip circumference were measured after voiding. Height was measured using a wall-mounted stadiometer, and body weight was measured to the nearest 0.1 kg using a calibrated scale. Waist circumference was measured at the mid-axillary line (midpoint between the lowest rib and the iliac crest), and hip circumference at the widest point of the buttocks. Body mass index was calculated as weight divided by height squared (kg/m^2^). Body composition (muscle mass, fat mass, fat-free mass, and total body water) was assessed by bioelectrical impedance analysis (InBody Co). Resting blood pressure was measured using an automated digital monitor after 10 minutes of seated rest. Grip strength was assessed by handgrip dynamometry using the dominant hand.

#### Autophagic flux measurement

Autophagic flux was assessed using methodology developed by our group (22). Briefly, 6 mL of blood was divided into 2 tubes – one served as the control, while the other was treated with 150 μM chloroquine (Sigma Aldrich, C6628), a lysosomal inhibitor. Samples were incubated for 1 h at 37°C with rotation. All subsequent steps were performed on ice or at 4°C to inhibit further vesicle trafficking. PBMCs were isolated, snap-frozen on dry ice and stored at −80°C for subsequent biochemical analysis.

PBMC pellets were thawed on ice and resuspended in PBS containing 0.05% saponin (Sigma Aldrich, SAE0073) for 5 min to selectively remove the cytosolic pool of LC3B-I, while preserving membrane-associated LC3B-II (19, 25). PBMCs were washed, lysed in cell extraction buffer (Cell Signalling Technology, 35172) containing protease inhibitors, and sonicated. Samples were centrifuged at 16,000 x g for 5 min at 4°C, with protein quantification performed on the clarified lysate.

LC3B-II concentration was measured by loading 5 μg clarified cell lysate per well in triplicate on a FastScan Total LC3B ELISA kit (Cell Signalling Technology, 35172). A standard curve was prepared using 0 - 4 ng/well recombinant human LC3B (Abcam, ab103506). ELISA plates were prepared as per manufacturer’s instructions, and the absorbance measured at 450 nm using Glomax plate reader (Promega). LC3B-II protein concentrations of each sample were interpolated from the standard curve and reported as ng LC3B-II/mg of total protein/hour. Autophagic flux was calculated as: ΔLC3B-II (ng LC3B-II/mg protein/hour) = LC3B-II [chloroquine] – LC3B-II [control]. If LC3B-II [control] was below the limit of detection, a value of 10.157 was used in the calculation (half of the lowest detected control value).

#### Blood based measurements

Plasma samples were isolated by centrifugation, stored at −80°C, and thawed immediately prior to analysis. Plasma insulin was measured in duplicate using a commercial ELISA (Mercodia, 10-1113-10) according to the manufacturer’s instructions. Concentrations were derived from a standard curve and converted from mU/L to pmol/L using a conversion factor of 6. Plasma glucose was measured using the hexokinase method on a Cobas Integra 400 plus analyser (Roche). Plasma triglycerides, total cholesterol, and high-density lipoprotein (HDL) cholesterol were measured on the same platform using standard enzymatic assays. Low-density lipoprotein (LDL) cholesterol was calculated using the Friedewald equation: LDL (mmol/L) = total cholesterol − HDL − (triglycerides/2.2).

#### Questionnaires

To assess the effects of dietary protein intake on sleep quality, eating behaviours, quality of life, and physical activity, participants completed a series of validated questionnaires at each study visit. These included the Short Form-36 (SF-36) to assess health-related quality of life (26), the Pittsburgh Sleep Quality Index (PSQI) to assess self-rated sleep quality (27), the Three-Factor Eating Questionnaire (TFEQ) to assess cognitive restraint, disinhibition, and hunger (28), and the International Physical Activity Questionnaire (IPAQ) (29). In addition, participants completed a brief survey at the end of each dietary intervention period to capture perceptions, expectations, and experiences related to the study diets.

#### Metabolomics

To explore broader metabolic adaptations, we performed metabolite profiling on fasting plasma collected at the end of each dietary intervention period. Briefly, 20 μL plasma was extracted in 140 μL of chilled 1:3:3 MilliQ water:acetonitrile:methanol containing isotopically labelled internal standards. Samples were vortexed and centrifuged, and 20 μL of the supernatant was dried under vacuum at 35°C. Dried extracts were reconstituted in methanol, dried again, and derivatised online using methoxyamine followed by BSTFA with 1% TMCS. Analyses were performed on a GC-triple quadrupole mass spectrometry platform. Study samples were randomized, internal standards were included in every sample, and pooled biological quality controls, pooled quality controls, and blanks were injected at regular intervals to monitor analytical performance. Peak areas were normalised to internal standards, log2-transformed and analysed using a paired linear model with participant as a blocking factor to estimate the effect of reduced-versus average-protein intake within individuals. Raw p values were adjusted for multiple comparisons using the Benjamini-Hochberg false discovery rate. Analyses were performed using R (version 4.4.2) and the limma package (30).

### Sample size

To detect a 15% within-subject difference in autophagic flux (ΔLC3B-II) between dietary conditions using paired t-test, assuming a mean (within-subject SD) autophagic flux of 277.9 (90.7), a sample size of 53 participants would provide approximately 90% power. To allow for potential attrition and missing data, 74 participants were enrolled.

### Statistical analysis

Statistical analyses were conducted blinded, according to a pre-specified statistical analysis plan. The primary analysis compared autophagic flux following the reduced-protein and average protein diets using a linear mixed-effects model which included fixed effects for diet, period, sex and randomization sequence, a random effect for participant and unstructured residual covariance across periods. Fixed effects for the difference in period baseline measurements of the outcome and the interaction between the difference in period baselines and period were also included to adjust for baseline differences between participants and over periods (31). The model was fitted using restricted maximum likelihood estimation and included participants with both baseline measurements and the outcome measured after at least one period. Secondary outcomes were analysed using analogous models, with transformations applied where required to improve model fit. Analyses were performed on all available data under an intention-to-treat principle. A sensitivity analysis for the primary outcome was conducted to assess the impact of missing data and model assumptions. The sensitivity analysis jointly modelled the outcome and baseline measurements, using a linear mixed effects model with a first-order autoregressive correlation structure for the residuals and Kenward-Roger degrees of freedom (31). The sensitivity analysis model included all participants with at least one baseline or outcome measurement, including those lost to follow up.

Effect estimates are presented with 95% confidence intervals and two-sided p-values. All analyses were performed using R (version 4.4.3) and Stata (version 18.5).

## Results

### Trial participants

This report is based on data from 74 healthy adults enrolled in a single-site, single-blinded, randomized crossover trial. Each participant was randomized to receive both an average-protein and reduced-protein dietary intervention, with 36 participants randomized to the average-protein followed by reduced-protein sequence, and 38 to the reduced-protein followed by average-protein sequence. Sixty-three participants completed both diet periods and attended the post-intervention assessments, with 61 participants included in the primary outcome analysis (Figure 1). Inclusion in the primary outcome analysis required both baseline measurements, and at least one post-intervention LC3B flux measurement. All exclusions were applied without reference to diet allocation. Baseline participant characteristics were similar between the two randomization sequences (Table 1).

**Table 1.** Summary of participant demographic characteristics by randomized group. Data are presented as mean (SD).

| Characteristic | Sequence Average-Reduced<br>(N=36) | Sequence Reduced-Average<br>(N=38) | Total<br>(N=74) |
| --- | --- | --- | --- |
| Age (years) | 28.22 (6.14) | 30.61 (7.97) | 29.45 (7.19) |
| Sex: n (%) |  |  |  |
| Female | 22 (61%) | 22 (58%) | 44 (59%) |
| Male | 14 (39%) | 16 (42%) | 30 (41%) |
| BMI (kg/m<sup>2</sup>) | 23.42 (3.23) | 24.62 (3.06) | 24.04 (3.18) |

**Figure 1.**
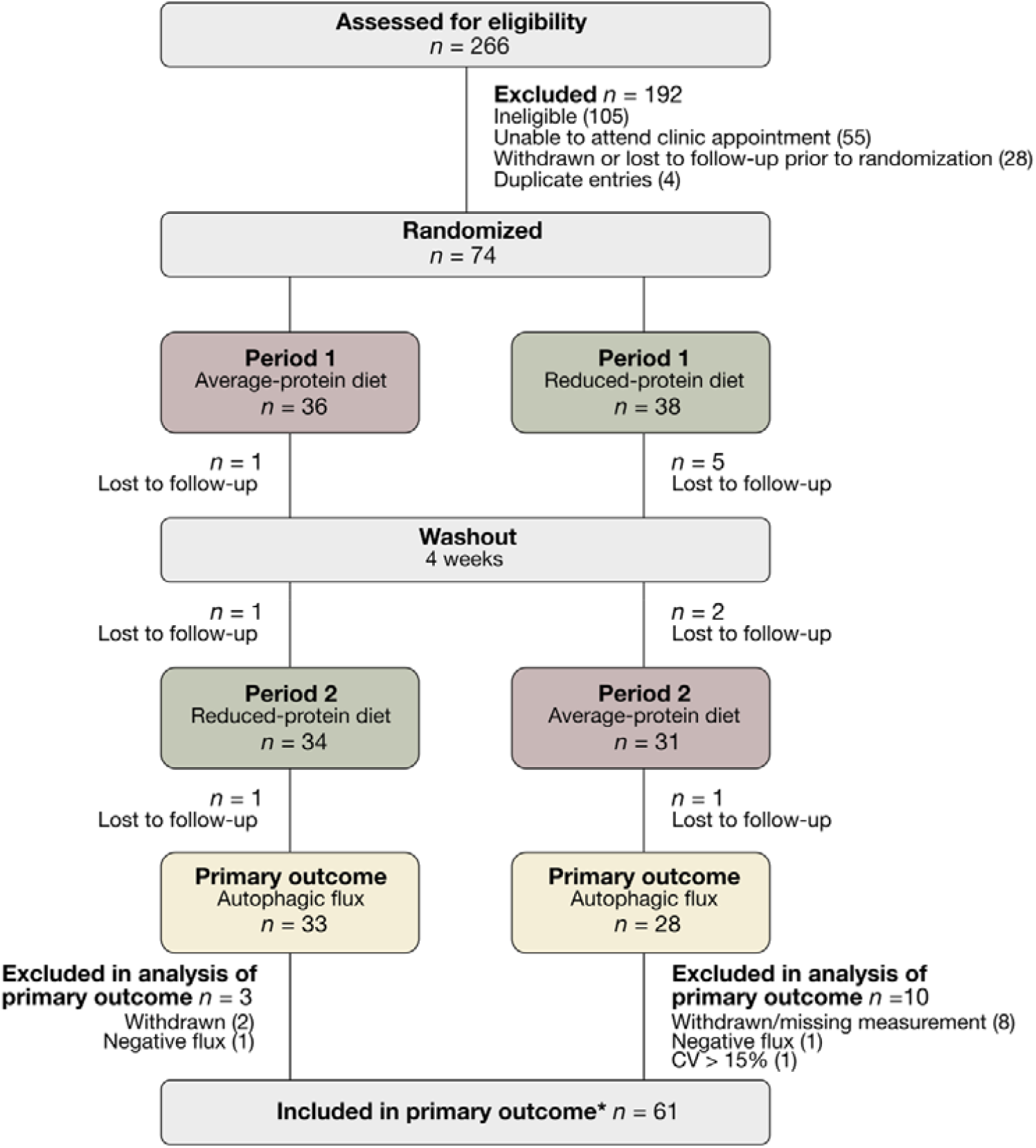
CONSORT Chart. Participants were randomly assigned to either the average-protein followed by reduced-protein sequence, or the reduced-protein followed by average-protein sequence. Each dietary intervention lasted 4 weeks and was separated by a 4-week washout period before crossover. *Primary outcome analysis required both baseline measurements and ≥ 1 outcome LC3 flux measurement. Of 11 participants lost to follow-up, 1 met the primary outcome analysis criteria and was included, and 10 were excluded following withdrawal before the required measurements were obtained.

### Autophagic flux

Mean baseline autophagic flux values were similar prior to each dietary intervention. There was no evidence that reducing dietary protein over a 4-week period altered autophagic flux. The primary analysis showed no difference in mean autophagic flux between the reduced- and average-protein interventions (adjusted mean difference: −8.46 ng LC3BII/mg protein/h; 95% CI: −24.06 to 7.14; p = 0.28; Table 2). The result of the sensitivity analysis was consistent with the primary analysis.

**Table 2.** Baseline and outcome measurements of autophagic flux by diet. Data are presented as mean (SD). *Adjusted for period, sex, randomization group (sequence), difference in period baselines and the interaction between the difference in period baselines and period. Estimated using linear mixed effect models of the two outcome measurements with a random subject effect for participant and unstructured residual covariance across periods, fitted using restricted maximum likelihood estimation. **Number of participants included in the adjusted analysis model.

|  | Average-Protein Diet |  | Reduced-Protein Diet |  |  |  |  |  |  |
| --- | --- | --- | --- | --- | --- | --- | --- | --- | --- |
| Variable | Baseline | Outcome | Baseline | Outcome | Diet effect estimate (Reduced - Average) Unadjusted | Diet effect estimate (Reduced - Average) Adjusted* | 95% confidence interval | p value | N included in the analysis** |
| Autophagic Flux (ng LC3BII/mg protein/h) | 200.68 (67.17)<br>n = 66 | 198.77 (57.21)<br>n = 62 | 200.33 (56.78)<br>n = 69 | 189.31 (65.75)<br>n = 64 | -7.18 (59.40)<br>n = 58 | -8.46 | (-24.06, 7.14) | 0.28 | 61 |

### Anthropometric and metabolic measures

Fasting glucose, insulin, triglycerides, total cholesterol, LDL cholesterol, blood pressure, and grip strength did not differ between the average- and reduced-protein dietary interventions (Table 3). HDL cholesterol was modestly higher following the reduced-protein diet compared with the average-protein diet. Small differences in body composition were observed between the reduced-protein and the average-protein interventions, including reductions in body weight, muscle mass, total body water, and fat-free mass, while fat mass was unchanged.

**Table 3.** Summary of metabolic and anthropometric secondary outcomes by diet. Data are presented as mean (SD). *Adjusted for period, sex, randomization group (sequence), difference in period baselines and the interaction between the difference in period baselines and period. Estimated using linear mixed effect models of the two outcome measurements with a random subject effect for participant and unstructured residual covariance across periods, fitted using restricted maximum likelihood estimation. **Number of participants included in the adjusted analysis model.

| Variable | Average-Protein Diet |  | Reduced-Protein Diet |  | Diet effect estimate<br>(Reduced - Average)<br>Unadjusted | Diet effect estimate<br>(Reduced - Average)<br>Adjusted* | 95% confidence interval | p value | N included in the analysis** |
| --- | --- | --- | --- | --- | --- | --- | --- | --- | --- |
|  | Baseline | Outcome | Baseline | Outcome |  |  |  |  |  |
| Glucose (mmol/L) | 5.00<br>(0.50)<br>n = 65 | 4.95<br>(0.50)<br>n = 60 | 4.98<br>(0.52)<br>n = 71 | 4.96<br>(0.51)<br>n = 63 | 0.02 (0.27)<br>n = 56 | 0.01 | (-0.06, 0.09) | 0.73 | 62 |
| Triglycerides (mmol/L) | 0.93<br>(0.39)<br>n = 66 | 0.88<br>(0.41)<br>n = 65 | 0.98<br>(0.53)<br>n = 72 | 0.93<br>(0.43)<br>n = 66 | 0.05 (0.33)<br>n = 63 | 0.06 | (-0.03, 0.14) | 0.18 | 64 |
| Total cholesterol (mmol/L) | 4.73<br>(0.76)<br>n = 66 | 4.47<br>(0.72)<br>n = 65 | 4.72<br>(0.77)<br>n = 72 | 4.53<br>(0.79)<br>n = 66 | 0.06 (0.54)<br>n = 63 | 0.05 | (-0.08, 0.18) | 0.45 | 64 |
| HDL (mmol/L) | 1.48<br>(0.33)<br>n = 66 | 1.40<br>(0.32)<br>n = 65 | 1.47<br>(0.34)<br>n = 72 | 1.45<br>(0.33)<br>n = 66 | 0.05 (0.16)<br>n = 63 | 0.05 | (0.01, 0.09) | 0.01 | 64 |
| LDL (mmol/L) | 2.83<br>(0.65)<br>n = 66 | 2.67<br>(0.64)<br>n = 65 | 2.81<br>(0.72)<br>n = 72 | 2.66<br>(0.70)<br>n = 66 | -0.01 (0.48)<br>n = 63 | -0.02 | (-0.14, 0.10) | 0.76 | 64 |
| Insulin | 32.38<br>(19.54)<br>n = 65 | 29.45<br>(15.71)<br>n = 65 | 32.69<br>(21.36)<br>n = 71 | 29.89<br>(17.92)<br>n = 66 | 0.35 (16.64)<br>n = 63 | 0.33 | (-3.90, 4.56) | 0.88 | 64 |
| Body weight (kg) | 66.71<br>(12.05)<br>n = 67 | 66.09<br>(12.12)<br>n = 65 | 66.66<br>(12.09)<br>n = 72 | 65.63<br>(11.89)<br>n = 66 | -0.30 (1.53)<br>n = 63 | -0.38 | (-0.68, -0.09) | 0.01 | 65 |
| Muscle mass (kg) | 27.15<br>(6.63)<br>n = 67 | 27.01<br>(6.45)<br>n = 64 | 27.01<br>(6.61)<br>n = 72 | 26.60<br>(6.50)<br>n = 66 | -0.29 (0.72)<br>n = 62 | -0.31 | (-0.47, -0.15) | <0.01 | 65 |
| Body fat mass (kg) | 18.00<br>(6.67)<br>n = 67 | 17.47<br>(6.56)<br>n = 65 | 18.18<br>(6.89)<br>n = 72 | 17.81<br>(6.57)<br>n = 66 | 0.18 (1.28)<br>n = 63 | 0.13 | (-0.14, 0.40) | 0.34 | 65 |
| Total body water (litres) | 35.67<br>(7.87)<br>n = 67 | 35.61<br>(7.66)<br>n = 65 | 35.50<br>(7.85)<br>n = 72 | 35.02<br>(7.75)<br>n = 66 | -0.36 (0.90)<br>n = 63 | -0.37 | (-0.57, -0.17) | <0.01 | 65 |
| Fat free mass (kg) | 48.70<br>(10.76)<br>n = 67 | 48.62<br>(10.44)<br>n = 65 | 48.48<br>(10.70)<br>n = 72 | 47.82<br>(10.57)<br>n = 66 | -0.48 (1.21)<br>n = 63 | -0.50 | (-0.77, -0.23) | <0.01 | 65 |
| Systolic blood | 113.84<br>(12.08) | 115.35<br>(12.44) | 114.93<br>(11.27) | 114.21<br>(11.90) | -0.52 (9.37) | -0.90 | (-3.18, | 0.43 | 65 |

| Variable | Average-Protein Diet |  | Reduced-Protein Diet |  | Diet effect estimate (Reduced - Average) Unadjusted | Diet effect estimate (Reduced - Average) Adjusted* | 95% confidence interval | p value | N included in the analysis** |
| --- | --- | --- | --- | --- | --- | --- | --- | --- | --- |
|  | Baseline | Outcome | Baseline | Outcome |  |  |  |  |  |
| pressure (mmHg) | n = 67 | n = 65 | n = 72 | n = 66 | n = 63 |  | 1.38) |  |  |
| Diastolic blood pressure (mmHg) | 73.76 (5.92)<br>n = 67 | 73.52 (5.51)<br>n = 65 | 74.35 (6.65)<br>n = 72 | 73.53 (6.22)<br>n = 66 | 0.29 (5.29)<br>n = 63 | 0.43 | (-0.88, 1.74) | 0.52 | 65 |
| Muscular grip strength (kg) | 35.60 (11.51)<br>n = 67 | 36.65 (11.56)<br>n = 65 | 35.75 (12.26)<br>n = 72 | 36.02 (11.68)<br>n = 66 | -0.29 (4.04)<br>n = 63 | -0.29 | (-1.32, 0.74) | 0.58 | 65 |

### Self-reported health, lifestyle and behavioural outcomes

Self-reported physical and mental health, sleep quality, eating behaviour, and physical activity did not differ between the average- and reduced-protein dietary interventions (Table 4). Scores across SF-36 subscales, Physical and Mental Component Scores, Pittsburgh Sleep Quality Index, eating behaviour questionnaires, and self-reported physical activity were broadly similar between diets. There were no significant changes in any self-reported outcome following the reduced-protein intervention.

**Table 4.** Summary of self-reported secondary outcomes by diet. Data are presented as mean (SD). *Adjusted for period, sex, randomization group (sequence), difference in period baselines and the interaction between the difference in period baselines and period. Estimated using linear mixed effect models of the two outcome measurements with a random subject effect for participant and unstructured residual covariance across periods, fitted using restricted maximum likelihood estimation. **IPAQ metabolic equivalent of task (MET) minutes/week was modelled using a log transform, so the adjusted effect estimate (95% CI) is the multiplicative effect on mean IPAQ MET minutes/week following the reduced-protein diet compared to the average-protein diet.

| Variable | Average-Protein Diet |  | Reduced-Protein Diet |  |  |  |  |  |  |
| --- | --- | --- | --- | --- | --- | --- | --- | --- | --- |
|  | Baseline | Outcome | Baseline | Outcome | Diet effect estimate (Reduced - Average) Unadjusted | Diet effect estimate (Reduced - Average) Adjusted* | 95% confidence interval | p value | N included in the analysis |
| SF-36 Mental Component Score | 45.14 (12.22)<br>n = 67 | 46.78 (11.98)<br>n = 65 | 44.83 (13.53)<br>n = 72 | 47.28 (10.56)<br>n = 66 | 0.43 (9.86)<br>n = 63 | 0.29 | (-2.00, 2.57) | 0.80 | 65 |
| SF-36 Physical Component Score | 56.52 (4.73)<br>n = 67 | 55.79 (6.24)<br>n = 65 | 55.78 (6.67)<br>n = 72 | 55.69 (4.82)<br>n = 66 | -0.43 (5.29)<br>n = 63 | -0.49 | (-1.85, 0.87) | 0.47 | 65 |
| Self-rated sleep habits (Pittsburgh Sleep Quality Index) | 5.52 (2.57)<br>n = 67 | 5.17 (2.72)<br>n = 65 | 5.74 (2.78)<br>n = 72 | 5.14 (2.97)<br>n = 66 | -0.13 (2.26)<br>n = 63 | -0.10 | (-0.67, 0.47) | 0.73 | 65 |
| Cognitive Restraint Scale Score | 14.90 (4.24)<br>n = 67 | 15.26 (4.38)<br>n = 65 | 14.81 (4.24)<br>n = 72 | 14.82 (4.27)<br>n = 66 | -0.37 (2.52)<br>n = 63 | -0.39 | (-1.02, 0.24) | 0.22 | 65 |
| Uncontrolled Eating Scale Score | 16.96 (4.84)<br>n = 67 | 17.55 (5.22)<br>n = 65 | 17.47 (4.67)<br>n = 72 | 16.88 (4.88)<br>n = 66 | -0.56 (2.59)<br>n = 63 | -0.57 | (-1.20, 0.05) | 0.07 | 65 |
| Emotional Eating Scale Score | 5.52 (2.05)<br>n = 67 | 5.52 (2.31)<br>n = 65 | 5.40 (2.21)<br>n = 72 | 5.76 (2.23)<br>n = 66 | 0.21 (1.44)<br>n = 63 | 0.28 | (-0.07, 0.62) | 0.11 | 65 |
| Physical activity (MET minutes/week according to IPAQ) | 3,397.24 (4,190.70)<br>n = 67 | 4,050.44 (5,254.49)<br>n = 65 | 3,413.60 (3,543.97)<br>n = 72 | 3,948.90 (5,346.47)<br>n = 66 | -37.34 (4599.23)<br>n = 63 | 0.97** | (0.74, 1.26)** | 0.80 | 65 |

### Exploratory metabolomic profiling

We conducted plasma metabolomic profiling at the end of each dietary intervention to assess metabolic responses to reduced protein intake. The reduced-protein diet induced changes in a small subset of metabolites (Figure 2, Supplementary Table 1). The higher circulating polyols – erythritol and threitol – observed following the reduced-protein diet reflect differences in the composition of the dietary supplement pouches used between interventions. Plasma urea was lower following the reduced-protein diet compared with the average-protein diet, consistent with reduced nitrogen disposal. Dihydroxyacetone was also lower following reduced protein intake. A small elevation in some circulating amino acids including glutamine, glycine, ornithine, and isoleucine was observed in the reduced-protein diet. Despite these diet-associated shifts, most metabolites were unchanged between diets. Effect sizes were generally small, with no broad suppression of amino acid pools observed.

**Figure 2.**
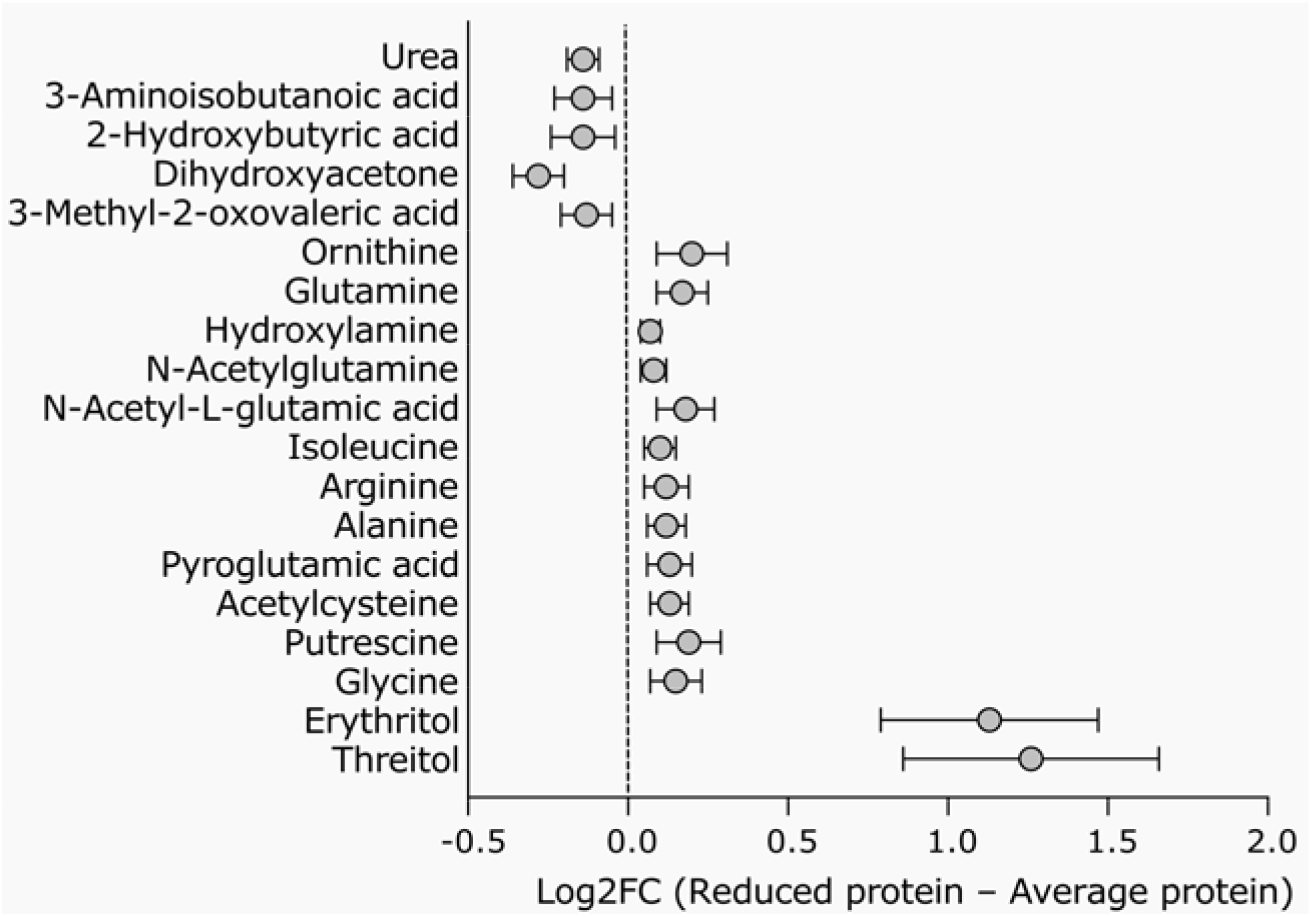
Plasma metabolites altered following reduced-protein intake. Estimated log2 fold-change (reduced-protein – average-protein diet) for the top plasma metabolites that differed significantly between dietary interventions.

## Discussion

In this randomized crossover trial conducted in healthy adults, we show that a 4-week reduction in dietary protein intake from 20% to 10% of energy did not alter fasting autophagic flux in human PBMCs. While nutrient restriction is widely viewed as a robust modulator of autophagy in animal models, our results add to a growing body of human data suggesting that autophagy regulation in circulating immune cells is more context-dependent (19, 32, 33) and may not mirror the canonical nutrient-mTORC1-autophagy coupling observed in tissues such as liver and skeletal muscle (12, 34–37).

Modifying nutrient intake impacts mTORC1 activity, leading to subsequent modulation of autophagy in preclinical models (34–36). In contrast, human evidence remains limited, primarily relying on static gene or protein markers (38, 39) which are known not to correlate well with autophagic flux, *i*.*e*. actual degradative capacity (8, 40), or measurements performed after refeeding cells in nutrient-rich culture medium (41) – conditions that can rapidly re-activate mTORC1 and obscure *in vivo* nutrient effects. Using a whole-blood assay that preserves the extracellular environment of PBMCs during lysosomal inhibition (22), we observed no meaningful difference in autophagic flux between reduced- and average-protein intake, suggesting that, at least in healthy adults, reducing dietary protein within recommended ranges does not translate to a detectable increase in autophagy in this cell population. Previously, we have shown that a single high-protein meal does not alter autophagic flux in human PBMCs despite changes in postprandial insulin, glucose and mTORC1 activity (19). Our present work tested whether a more sustained dietary adaptation over four weeks might produce effects not apparent in the short-term. However, we found that 4-week protein reduction similarly failed to modulate basal autophagy in circulating immune cells.

A previous report showed that one year of a low protein diet in patients with collagen VI-related myopathies increased transcript and protein levels of LC3B and BECN1 in muscle and leukocytes (42), although the sample size was small and flux was not examined. This suggests that a longer intervention period may demonstrate changes in autophagy. Recently, another group reported an increase in some autophagy transcripts including LC3A, an LC3B paralog, in people after 7 days of a low-protein fasting mimicking diet (FMD) compared to both a high-protein FMD and a control diet (39). However, they did not observe concomitant increases in protein levels of any of the autophagy genes measured. Further, changes in autophagy observed by Burns et al (39) could be driven primarily by the FMD rather than modulation of dietary protein intake per se. Consistent with this interpretation, a separate study recently demonstrated increased conversion of LC3B to its lipidated form in people after 5 days of FMD without manipulation of dietary protein content (43), albeit they did not observe any differences in autophagic flux between control and FMD.

Previous work has shown a decrease in monocyte autophagy after a high protein meal (15), although the study employed LC3 immunofluorescence rather than flux measurements. This is in line with our previous work showing that in the human PBMC pool, monocytes – in particular, non-classical monocytes – are the only subpopulation that responded to amino acid restriction by increasing their autophagy (33). In this present study, flux was assessed in the bulk PBMC pool, and not at a cell type-specific resolution. Therefore, our null finding does not necessarily rule out cell type-restricted effects, a limitation of our work. Further, blood sampling was performed in the fasted state only. If reduced protein intake alters PBMC autophagy transiently during postprandial windows or under acute stress, our design may not have captured such effects. A further consideration is that, in contrast to other tissues, PBMC autophagy may be relatively buffered against modest changes in macronutrient composition and therefore may not reflect changes in autophagy that occur in other tissues. Indeed, in an aging mouse model of autophagy, we have observed that changes in PBMC autophagy do not correspond with that of tissues such as the brain and heart (6). This represents an inherent limitation of human autophagy measurement as blood is, at present, the most accessible source of human tissue amenable to flux assays. Using our methodology, we and others have previously observed an increase in PBMC autophagic flux with aging (5, 8) and intermittent time-restricted eating (25), and decreased flux following the addition of exogenous insulin and leucine to blood (22). This indicates that, at least in some scenarios, our assay can detect changes to autophagy.

The reduced-protein intervention achieved effective dietary separation, as confirmed by lower circulating urea, consistent with reduced nitrogen disposal and altered amino acid handling (44). We also observed minor differences in body composition compared with the average-protein intervention, including small reductions in body weight, fat-free mass, muscle mass and total body water, and a modest attenuation in the decline of HDL cholesterol. However, the overall metabolomic and physiological response was limited, with most amino acids and metabolites unchanged and no differences in self-reported health status, sleep quality, eating behaviours, physical activity or quality of life. This indicates that short-term protein reduction within guideline ranges is well-tolerated in healthy adults and suggests that our null autophagy result likely cannot be attributed to masking by compensatory stress responses.

In summary, our findings indicate that reducing dietary protein intake for four weeks does not increase basal autophagic flux in circulating immune cells of healthy adults. These data suggest that protein restriction alone, in the absence of caloric deficit or additional metabolic stressors, is unlikely to be a robust strategy for activating autophagy in human blood. Defining which interventions can demonstrably modulate human autophagy is a necessary prerequisite for rationally designing strategies to target age-related autophagy dysfunction. Future studies should examine whether more pronounced dietary interventions, longer durations, or combinations with other autophagy-inducing stimuli are required to modulate autophagic flux in humans, and whether responses differ across tissues, age groups, or metabolic states.

## Data Availability

All data produced in the present study are available upon reasonable request to the authors

## Abbreviations

mTORC1: mechanistic target of rapamycin 1
PBMC: peripheral blood mononuclear cells;
LC3: microtubule-associated protein 1A/1B light chain 3B;
BMI: body mass index;
HDL: High-density lipoprotein
LDL: Low-density lipoprotein
ELISA: enzyme-linked immunosorbent assay
PSQI: Pittsburgh Sleep Quality Index
TFEQ: Three-Factor Eating Questionnaire
IPAQ: International Physical Activity Questionnaire
MET: Metabolic equivalent of task
SF-36: Short Form-36
HDL: high-density lipoprotein
LDL: low-density lipoprotein
FMD: fasting-mimicking diet

## Data availability

De-identified individual participant data, statistical code, the statistical analysis plan and other study materials supporting the findings of this study are available from the corresponding author upon reasonable request.

## Acknowledgements

The authors would like to express their gratitude to the donors of the Standard Award Program in Alzheimer’s Disease Research grant (A2021040S), a program of the BrightFocus Foundation, for support of this research. We acknowledge A/Prof Lisa Yelland for advising on the statistical aspects of this study, the University of Adelaide HREC for reviewing our ethics application, and Dr Brunda Nijagal from the Metabolomics Australia at the University of Melbourne for performing the plasma metabolomics analysis.

## Author contribution

Conceptualization: TJS, CF, JB, LKH, XTT and SS conceived the study. LKH and XTT designed the dietary interventions. Funding acquisition: TJS, JB, LKH obtained funding for the study. Project administration: JRG assisted with writing the protocol and essential documents, obtained ethics approval, registered the study and was responsible for overall project management. and registered the study. GB conducted participant visits, coordinated food deliveries and maintained study records. Investigation: LKH, KJH, BK and AM performed the assays used in this study. Data analysis: SS, KB, KL and LY analysed the data generated in this study. Writing: SS and TJS wrote the first draft of the manuscript. Review and editing: All authors contributed critical subject matter expertise, reviewed and approved the final version of the manuscript.

